# Comparative effectiveness of BNT162b2 versus mRNA-1273 boosting in England: a cohort study in OpenSAFELY-TPP

**DOI:** 10.1101/2022.07.29.22278186

**Authors:** William J Hulme, Elsie MF Horne, Edward PK Parker, Ruth H Keogh, Elizabeth J Williamson, Venexia Walker, Tom Palmer, Helen J Curtis, Alex Walker, Amir Mehrkar, Jessica Morley, Brian MacKenna, Sebastian CJ Bacon, Ben Goldacre, Miguel A Hernán, Jonathan AC Sterne, the OpenSAFELY collaborative

## Abstract

**Introduction:** The COVID-19 booster vaccination programme in England used both BNT162b2 and mRNA-1273 vaccines. Direct comparisons of the effectiveness against severe COVID-19 of these two vaccines for boosting have not been made in trials or observational data.

**Methods:** On behalf of NHS England, we used the OpenSAFELY-TPP database to match adult recipients of each vaccine type on date of vaccination, primary vaccine course, age, and other characteristics. Recipients were eligible if boosted between 29 October 2021 and 31 January 2022, and followed up for 12 weeks. Outcomes were positive SARS-CoV-2 test, COVID-19 hospitalisation, and COVID-19 death. We estimated the cumulative incidence of each outcome, and quantified comparative effectiveness using risk differences (RD) and hazard ratios (HRs).

**Results:** 1,528,431 people were matched in each group, contributing a total 23,150,504 person-weeks of follow-up. The 12-week risks per 1,000 people of positive SARS-CoV-2 test were 103.2 (95%CI 102.4 to 104.0) for BNT162b2 and 96.0 (95.2 to 96.8) for mRNA-1273: the HR comparing mRNA-1273 with BNT162b2 was 0.92 (95%CI 0.91 to 0.92). For COVID-19 hospitalisations the 12-week risks per 1,000 were 0.65 (95%CI 0.56 to 0.75) and 0.44 (0.36 to 0.54): HR 0.67 (95%CI 0.58 to 0.78). COVID-19 deaths were rare: the 12-week risks per 1,000 were 0.03 (95%CI 0.02 to 0.06) and 0.01 (0.01 to 0.02): HR 1.23 (95%CI 0.59 to 2.56). Comparative effectiveness was generally similar within subgroups defined by the primary course vaccine brand, age, prior SARS-CoV-2 infection and clinical vulnerability.

**Conclusion:** Booster vaccination with mRNA-1273 COVID-19 vaccine was more effective than BNT162b2 in preventing SARS-CoV-2 infection and COVID-19 hospitalisation during the first 12 weeks after vaccination, during a period of Delta followed by Omicron variant dominance.

## Background

The UK COVID-19 vaccination programme delivered its first “booster” (third) doses in September 2021 (1). Based on guidance from the Joint Committee for Vaccination and Immunisation (JCVI) (2), booster vacciantion was initially offered to groups at high risk of severe COVID-19 disease, then progressively extended to the whole adult population by mid December 2021 (3) (4) (5). The BNT162b2 Pfizer-BioNTech vaccine was used initially, with a half-dose of mRNA-1273 Moderna vaccine also used from 29 October 2021 onwards. Concurrent boosting with BNT162b2 and mRNA-1273, receipt of which was largely determined by local availability rather than clinical criteria, enables a direct comparison of their effectiveness against SARS-CoV-2 positive tests and severe COVID-19 disease. No randomised trials have made such a comparison.

On behalf of NHS England, we used the OpenSAFELY-TPP database, covering 40% of English primary care practices and linked to national coronavirus surveillance, hospital episodes, and death registry data, to compare the effectiveness of boosting with BNT162b2 and mRNA-1273 in adults. Follow up encompassed the 29 October 2021 until 31 January 2022, a period of Delta then Omicron variant dominance.

## Methods

### Data source

All data were linked, stored and analysed securely within the OpenSAFELY platform: https://opensafely.org/. With the approval of NHS England, primary care records managed by the GP software provider TPP were linked, using NHS numbers, to A&E attendance and in-patient hospital spell records via NHS Digital’s Hospital Episode Statistics (HES), national coronavirus testing records via the Second Generation Surveillance System (SGSS), and national death registry records from the Office for National Statistics (ONS). COVID-19 vaccination history and health and social care worker status is available in the GP record directly via the National Immunisation Management System (NIMS).

### Eligibility criteria

We considered all adults (aged 18 years and over) who received a booster dose of BNT162b2 or mRNA-1273 between 29 October 2021 and 31 January 2022 inclusive, during which time both vaccine brands were being used. People were eligible if they: were registered at a GP practice using TPP’s SystmOne clinical information system at the time of boosting; received a two-dose primary vaccination course of either BNT162b2 or ChAdOx1-S (mixed dosing and mRNA-1273 were not considered due to small numbers); were not a health or social care worker, not resident in a care or nursing home and not medically housebound or receiving end-of-life care; had no evidence of SARS-CoV-2 infection or COVID-19 disease within the previous 90 days; were not hospitalised at the time of boosting; and had complete information on sex, deprivation, and Sustainability and Transformation Partnership (STP, a geographical grouping of NHS and local authorities).

### Matching

BNT162b2 and mRNA-1273 booster recipients were matched 1-1 without replacement, on the following characteristics: date of booster dose; primary vaccine course (BNT162b2 or ChadOx1-S); date of second vaccine dose (7 day caliper); sex (male or female); age (3 year caliper and within age groups defined by JVCI risk groups); clinical risk group defined by JCVI (clinically extremely vulnerable, clinically at-risk, neither); Index of Multiple Deprivation (IMD, grouped by quintile); STP as a surrogate for geographical region; evidence of prior SARS-CoV-2 infection (any of positive SARS-CoV-2 test, probable infection documented in primary care, or COVID-19 hospital attendance or admission); and morbidity count (0, 1, or 2 or more of diabetes, BMI over 40kg/m^2^, chronic heart disease, chronic kidney disease, chronic liver disease, chronic respiratory disease or severe asthma, chronic neurological disease, cancer within 3 years). The supplementary materials provide more information on how these characteristics were defined.

### Outcomes

We considered three outcomes. Positive SARS-CoV-2 tests were identified using SGSS testing records and based on swab date. Both polymerase chain reaction (PCR) and lateral flow test results were included, without differentiation between symptomatic and asymptomatic infection. COVID-19 hospitalisation was identified using HES in-patient hospital records with International Statistical Classification of Diseases and Related Health Problems 10th Revision (ICD-10) codes U07.1 or U07.2 as the reason for admission (6). COVID-19 death was defined as death with U07.1 or U07.2 ICD-10 codes mentioned anywhere on the death certificate (i.e., as an underlying or contributing cause of death).

### Statistical Analysis

Each person was followed from the day of boosting (time zero) for 12 weeks, or until the earliest of the outcome of interest, death, practice de-registration, or 31 January 2022.

Baseline characteristics for each vaccine group were tabulated, with between-group balance examined using standardised mean differences. We estimated the cumulative incidence of each outcome in each vaccine group using the Kaplan-Meier (KM) estimator. We estimated risk differences (RDs) and hazard ratios (HRs) comparing the vaccine groups for each outcome. We used Cox models to estimate hazard ratios (HRs) comparing vaccine groups.

95% confidence limits for the cumulative incidences were derived using standard errors on the log-KM scale. Confidence limits for the risk differences were derived from the sum of squares of the standard errors on the KM scale, using Greenwood’s formula. Confidence limits for the risk ratio were derived from the sum of squares of the standard errors on the log-KM scale.

### Subgroup analyses

We estimated comparative effectiveness separately in the following subgroups: primary vaccine course (ChAdOx1-S or BNT162b2); age (18-64 or ≥ 65 years); clinical vulnerability, as defined by JCVI (not clinically at-risk, clinically at-risk, or clinically extremely vulnerable); evidence of prior SARS-CoV-2 infection or not. We used χ^2^ tests to examine evidence for heterogeneity in comparative effectiveness between subgroups.

### Disclosure control

To satisfy strict re-identification minimisation requirements for statistical outputs from OpenSAFELY’s Trusted Research Environment, plots of cumulative event counts and the Kaplan-Meier cumulative incidence estimates were rounded such that each increment is based on at least 6 events. Event rates, risk differences, and risk ratios were derived from these rounded estimates.

### Software, code, and reproducibility

Data management and analyses were conducted using OpenSAFELY tools, Python version 3.8.10 and R version 4.0.2. All code is shared openly for review and re-use under MIT open license at https://github.com/opensafely/comparative-booster. Codelists are available at https://www.opencodelists.org/. The supplementary materials provide further details of codelists and data sources used for all variables in the study. Detailed pseudonymised patient data is potentially re-identifiable and therefore not shared.

## Results

### Study population and matching

10,139,268 adults registered at a TPP practice received a booster vaccination with either BNT162b2 (7,239,423) or mRNA-1273 (2,899,845) during the study period, with 6,264,525 (86.5%) and 6,264,525 (87.1%) eligible for matching (Figure 1). Eligible BNT162b2 booster recipients were on average older, more deprived and had higher rates of prior clinical conditions than mRNA-1273 booster recipients (Table 1). They were also more likely to have received BNT162b2 as their primary vaccination course (38% versus 30%).

**Figure 1:**
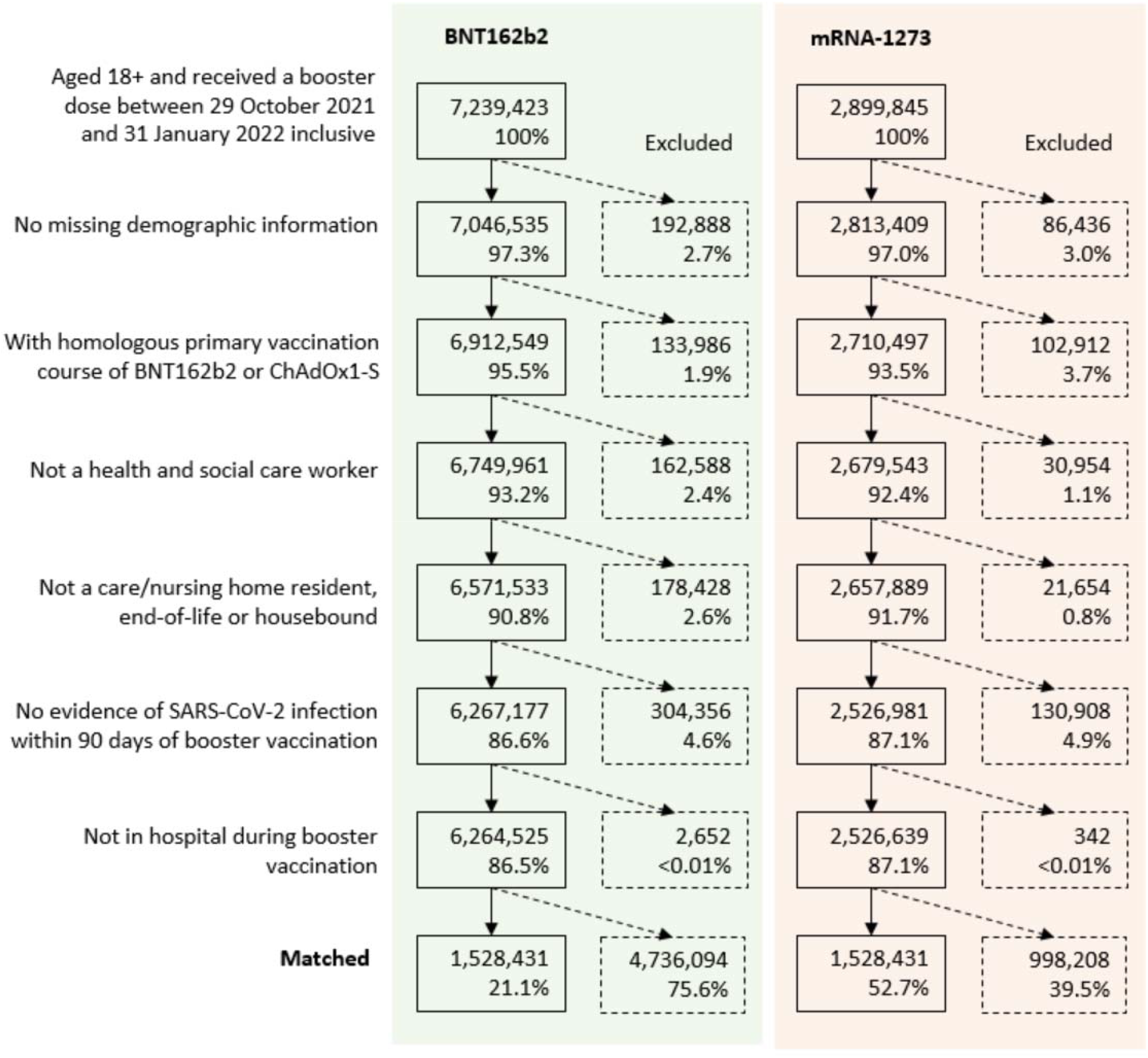
Flow of participants into the study

**Figure 2:**
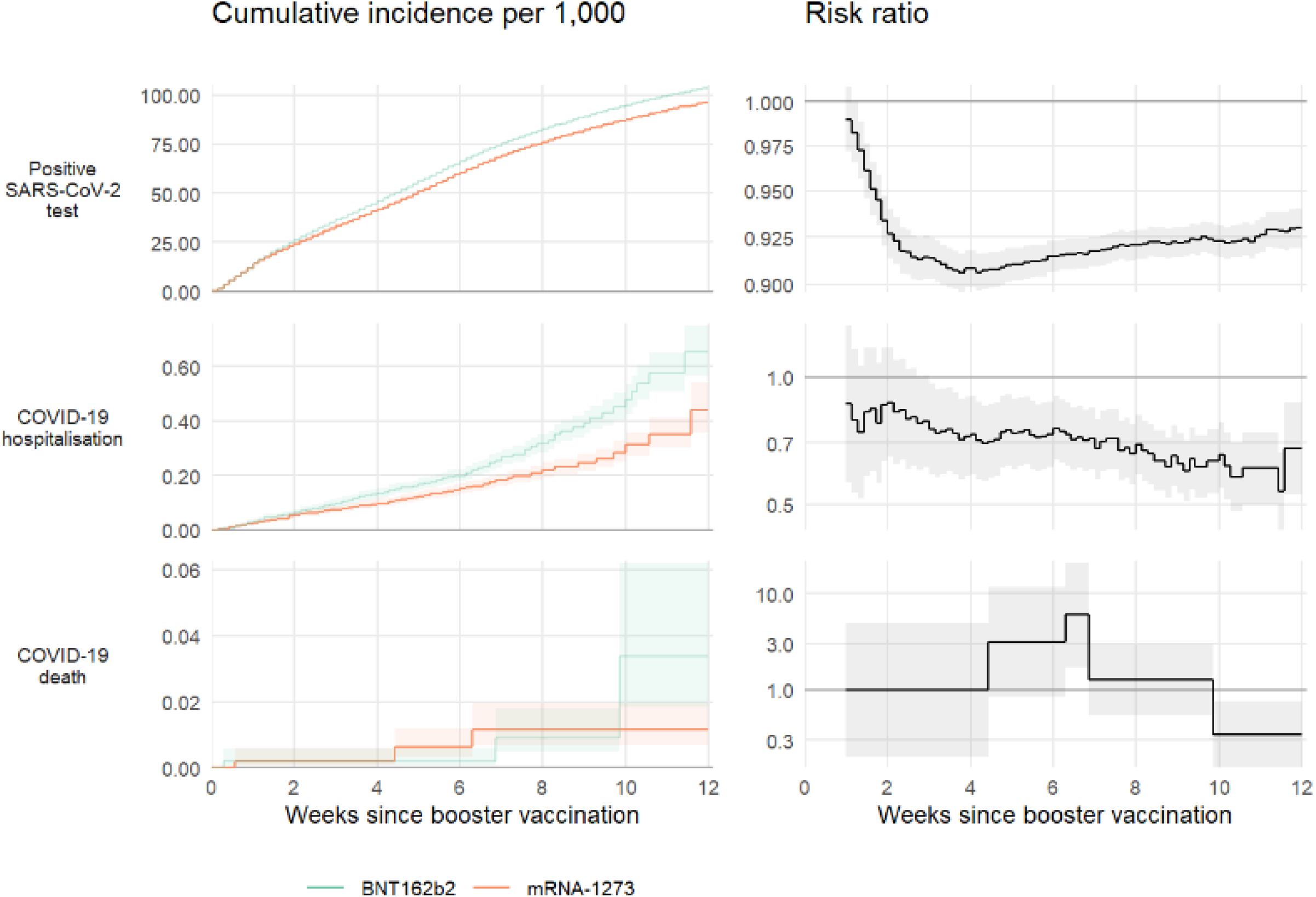
Estimates of cumulative risk per 1,000 people (left plots) and risk ratios (right plots) during the 12 weeks of follow up.

**Table 1:** Baseline characteristics before and after matching

| Variable | Characteristic | Before matching |  | After matching |  |
| --- | --- | --- | --- | --- | --- |
|  |  | BNT162b2 | mRNA-1273 | BNT162b2 | mRNA-1273 |
| Total |  | 6,264,527 | 2,526,641 | 1,528,431 | 1,528,431 |
| Primary vaccine | BNT162b2 | 2,375,489 (38%) | 755,196 (30%) | 452,292 (30%) | 452,292 (30%) |
|  | ChAdOx1 | 3,889,038 (62%) | 1,771,445 (70%) | 1,076,139 (70%) | 1,076,139 (70%) |
| Age | 18-39 | 1,416,562 (23%) | 624,521 (25%) | 394,393 (26%) | 392,356 (26%) |
|  | 40-49 | 975,152 (16%) | 509,126 (20%) | 296,512 (19%) | 298,740 (20%) |
|  | 50-59 | 1,377,740 (22%) | 714,867 (28%) | 447,312 (29%) | 446,312 (29%) |
|  | 60-69 | 1,357,668 (22%) | 506,744 (20%) | 307,632 (20%) | 309,646 (20%) |
|  | 70-79 | 924,289 (15%) | 143,800 (5.7%) | 77,389 (5.1%) | 76,079 (5.0%) |
|  | 80-89 | 182,800 (2.9%) | 23,687 (0.9%) | 4,855 (0.3%) | 4,909 (0.3%) |
|  | 90+ | 30,316 (0.5%) | 3,896 (0.2%) | 338 (<0.1%) | 389 (<0.1%) |
| Sex | Female | 3,184,371 (51%) | 1,222,957 (48%) | 725,115 (47%) | 725,115 (47%) |
|  | Male | 3,080,156 (49%) | 1,303,684 (52%) | 803,316 (53%) | 803,316 (53%) |
| Ethnicity | White | 5,256,075 (84%) | 2,117,344 (84%) | 1,281,776 (84%) | 1,280,089 (84%) |
|  | Black | 79,069 (1.3%) | 28,781 (1.1%) | 14,098 (0.9%) | 13,808 (0.9%) |
|  | South Asian | 343,343 (5.5%) | 117,499 (4.7%) | 67,028 (4.4%) | 63,792 (4.2%) |
|  | Mixed | 52,449 (0.8%) | 21,568 (0.9%) | 12,149 (0.8%) | 12,256 (0.8%) |
|  | Other | 100,548 (1.6%) | 40,573 (1.6%) | 22,977 (1.5%) | 23,467 (1.5%) |
|  | Unknown | 433,043 (6.9%) | 200,876 (8.0%) | 130,403 (8.5%) | 135,019 (8.8%) |
| Deprivation | 1 (most deprived) | 977,404 (16%) | 353,362 (14%) | 188,147 (12%) | 188,147 (12%) |
|  | 2 | 1,167,414 (19%) | 449,944 (18%) | 264,080 (17%) | 264,080 (17%) |
|  | 3 | 1,411,881 (23%) | 557,366 (22%) | 350,832 (23%) | 350,832 (23%) |
|  | 4 | 1,394,475 (22%) | 575,680 (23%) | 365,384 (24%) | 365,384 (24%) |
|  | 5 (least deprived) | 1,313,353 (21%) | 590,289 (23%) | 359,988 (24%) | 359,988 (24%) |
| Region | North East and Yorkshire | 1,103,864 (18%) | 459,633 (18%) | 307,129 (20%) | 307,129 (20%) |
|  | North West | 494,916 (7.9%) | 292,083 (12%) | 153,432 (10%) | 153,432 (10%) |
|  | Midlands | 1,400,469 (22%) | 489,643 (19%) | 286,987 (19%) | 286,987 (19%) |
|  | East of England | 1,336,998 (21%) | 740,850 (29%) | 427,468 (28%) | 427,468 (28%) |
|  | London | 310,882 (5.0%) | 147,195 (5.8%) | 94,201 (6.2%) | 94,201 (6.2%) |
|  | South East | 525,406 (8.4%) | 85,115 (3.4%) | 56,637 (3.7%) | 56,637 (3.7%) |
|  | South West | 1,091,992 (17%) | 312,122 (12%) | 202,577 (13%) | 202,577 (13%) |
| JCVI risk group | Not at-risk | 4,325,858 (69%) | 1,984,460 (79%) | 1,333,677 (87%) | 1,333,677 (87%) |
|  | At-risk | 1,465,750 (23%) | 439,990 (17%) | 178,879 (12%) | 178,879 (12%) |
|  | Extremely vulnerable | 472,919 (7.5%) | 102,191 (4.0%) | 15,875 (1.0%) | 15,875 (1.0%) |
| Morbidities | BMI >40 kg/m<sup>2</sup> | 262,656 (4.2%) | 97,119 (3.8%) | 21,449 (1.4%) | 21,533 (1.4%) |
|  | Chronic heart disease | 684,780 (11%) | 173,075 (6.9%) | 69,412 (4.5%) | 70,330 (4.6%) |
|  | Chronic kidney disease | 265,999 (4.2%) | 50,250 (2.0%) | 19,427 (1.3%) | 19,270 (1.3%) |
|  | Diabetes | 647,619 (10%) | 178,885 (7.1%) | 62,846 (4.1%) | 62,675 (4.1%) |
|  | Chronic liver disease | 165,248 (2.6%) | 55,914 (2.2%) | 20,289 (1.3%) | 20,221 (1.3%) |
|  | Chronic respiratory disease | 255,545 (4.1%) | 59,020 (2.3%) | 19,706 (1.3%) | 19,520 (1.3%) |
|  | Asthma | 33,538 (0.5%) | 11,484 (0.5%) | 3,521 (0.2%) | 3,577 (0.2%) |
|  | Chronic neurological. disease | 327,345 (5.2%) | 93,361 (3.7%) | 34,474 (2.3%) | 33,925 (2.2%) |
|  | Cancer, previous 3 years | 151,287 (2.4%) | 36,679 (1.5%) | 10,141 (0.7%) | 9,889 (0.6%) |
|  | Immunosuppressed | 135,638 (2.2%) | 29,798 (1.2%) | 6,702 (0.4%) | 6,080 (0.4%) |
|  | Asplenia/poor spleen function | 46,435 (0.7%) | 14,027 (0.6%) | 2,679 (0.2%) | 2,690 (0.2%) |
|  | Learning disabilities | 42,424 (0.7%) | 12,176 (0.5%) | 2,892 (0.2%) | 2,072 (0.1%) |
|  | Serious mental illness | 62,596 (1.0%) | 21,195 (0.8%) | 4,677 (0.3%) | 4,217 (0.3%) |
| No. of SARS-CoV-2 tests in last 6 months | 0 | 3,317,613 (53%) | 1,228,452 (49%) | 766,844 (50%) | 739,152 (48%) |
|  | 1 | 1,128,354 (18%) | 478,774 (19%) | 290,908 (19%) | 291,489 (19%) |
|  | 2 | 547,697 (8.7%) | 243,026 (9.6%) | 143,451 (9.4%) | 147,451 (9.6%) |
|  | 3+ | 1,270,863 (20%) | 576,389 (23%) | 327,228 (21%) | 350,339 (23%) |
| Prior documented SARS-CoV-2 infection |  | 548,001 (8.7%) | 246,453 (9.8%) | 87,928 (5.8%) | 87,928 (5.8%) |

After matching, 1,528,431 people remained in each group and were included in analyses of comparative effectiveness, representing 24.4% and 60.5% of eligible BNT162b2 and mRNA-1273 recipients respectively (Figure 1, Figure S1). Characteristics were well-balanced between vaccine groups at the start of follow up (Table 1) with standardised mean differences consistently below 0.05 (supplementary Figure S2). In particular, prior clinical conditions, which were not directly matched on, were well-balanced between the groups after, though not before, matching.

### Estimated comparative effectiveness

There were 224,640 positive SARS-CoV-2 tests, 786 COVID-19 hospitalisations, and 30 COVID-19 deaths at 12-weeks across 23,150,504 person-weeks of follow-up (Table 2).

**Table 2:** Estimates of the 12-week risk per 1000 people in each vaccine group, and corresponding risk differences, overall and within subgroups

| Sub-group |  | BNT162b2 |  |  | mRNA-1273 |  |  | Risk difference per 1,000 (95% CI) |
| --- | --- | --- | --- | --- | --- | --- | --- | --- |
|  |  | Person-weeks | Events | 12-week risk per 1,000 (95% CI) | Person-weeks | Events | 12-week risk per 1,000 (95% CI) |  |
| Positive SARS-CoV-2 test |  |  |  |  |  |  |  |  |
| All |  | 11,144,667 | 117,045 | 103.2 (102.4 to 104.0) | 11,179,367 | 107,595 | 96.0 (95.2 to 96.8) | -7.24 (-8.37 to -6.11) |
| Primary vaccine course | ChAdOx1-S | 8,296,796 | 77,829 | 94.9 (94.0 to 95.8) | 8,318,683 | 72,183 | 89.5 (88.6 to 90.4) | -5.39 (-6.68 to -4.10) |
|  | BNT162b2 | 2,847,842 | 39,213 | 125.2 (123.5 to 127.0) | 2,860,668 | 35,415 | 113.5 (111.9 to 115.2) | -11.66 (-14.09 to -9.24) |
| Age | 18-64 | 9,138,153 | 110,943 | 123.9 (122.2 to 125.6) | 9,179,949 | 102,003 | 117.4 (115.5 to 119.3) | -6.53 (-9.04 to -4.02) |
|  | 65 and over | 2,006,490 | 6,099 | 36.7 (35.7 to 37.7) | 1,999,400 | 5,595 | 33.9 (33.0 to 34.9) | -2.75 (-4.11 to -1.39) |
| Prior SARS-CoV-2 infection | Yes | 10,554,634 | 112,455 | 104.6 (103.8 to 105.4) | 10,587,238 | 103,599 | 97.4 (96.6 to 98.2) | -7.21 (-8.36 to -6.06) |
|  | No | 590,008 | 4,587 | 73.9 (69.9 to 78.0) | 592,102 | 3,993 | 65.8 (62.6 to 69.1) | -8.09 (-13.26 to -2.91) |
| JCVI clinical risk group | Not at-risk | 9,365,204 | 105,969 | 109.1 (108.1 to 110.1) | 9,396,643 | 97,329 | 101.7 (100.7 to 102.7) | -7.44 (-8.86 to -6.02) |
|  | At-risk | 1,626,829 | 10,149 | 69.8 (68.3 to 71.3) | 1,629,565 | 9,453 | 65.4 (63.9 to 66.9) | -4.36 (-6.48 to -2.25) |
|  | Extremely vulnerable | 152,665 | 927 | 71.4 (66.8 to 76.2) | 153,184 | 813 | 62.3 (58.0 to 66.8) | -9.10 (-15.55 to -2.66) |
| COVID-19 hospitalisation |  |  |  |  |  |  |  |  |
| All |  | 11,573,743 | 471 | 0.65 (0.56 to 0.75) | 11,573,858 | 315 | 0.44 (0.36 to 0.54) | -0.21 (-0.34 to -0.08) |
| Primary vaccine course | ChAdOx1-S | 8,591,940 | 351 | 0.64 (0.53 to 0.76) | 8,592,440 | 243 | 0.58 (0.44 to 0.78) | -0.05 (-0.25 to 0.15) |
|  | BNT162b2 | 2,981,788 | 123 | 1.02 (0.75 to 1.40) | 2,981,397 | 75 | 0.38 (0.28 to 0.52) | -0.64 (-0.98 to -0.30) |
| Age | 18-64 | 9,542,708 | 303 | 0.45 (0.37 to 0.53) | 9,551,665 | 189 | 0.31 (0.23 to 0.42) | -0.14 (-0.26 to -0.01) |
|  | 65 and over | 2,031,006 | 165 | 1.00 (0.85 to 1.18) | 2,022,166 | 129 | 0.81 (0.67 to 0.98) | -0.19 (-0.42 to 0.03) |
| Prior SARS-CoV-2 infection | Yes | 10,967,752 | 459 | 0.75 (0.63 to 0.90) | 10,967,802 | 303 | 0.45 (0.36 to 0.55) | -0.30 (-0.47 to -0.14) |
|  | No | 605,960 | 15 | 0.29 (0.16 to 0.51) | 606,048 | 9 | 0.10 (0.05 to 0.20) | -0.19 (-0.36 to -0.01) |
| JCVI clinical risk group | Not at-risk | 9,751,542 | 249 | 0.34 (0.28 to 0.40) | 9,751,335 | 177 | 0.40 (0.27 to 0.60) | 0.07 (-0.11 to 0.24) |
|  | At-risk | 1,666,233 | 171 | 1.23 (1.04 to 1.44) | 1,666,339 | 105 | 0.70 (0.57 to 0.86) | -0.52 (-0.77 to -0.28) |
|  | Extremely vulnerable | 155,910 | 45 | 3.45 (2.56 to 4.66) | 156,122 | 33 | 2.52 (1.77 to 3.59) | -0.93 (-2.30 to 0.44) |
| COVID-19 death |  |  |  |  |  |  |  |  |
| All |  | 11,575,418 | 15 | 0.03 (0.02 to 0.06) | 11,575,086 | 15 | 0.01 (0.01 to 0.02) | -0.02 (-0.04 to 0.00) |
| Primary vaccine course | ChAdOx1-S | 8,593,205 | 9 | 0.01 (0.01 to 0.02) | 8,593,422 | 9 | 0.01 (0.00 to 0.02) | 0.00 (-0.01 to 0.01) |
|  | BNT162b2 | 2,982,189 | 3 | 0.04 (0.01 to 0.12) | 2,981,653 | 3 | 0.01 (0.00 to 0.02) | -0.03 (-0.08 to 0.01) |
| Age | 18-64 | 9,543,745 | 3 | 0.00 (0.00 to 0.01) | 9,552,357 | 3 | 0.00 (0.00 to 0.01) | 0.00 (0.00 to 0.00) |
|  | 65 and over | 2,031,640 | 9 | 0.06 (0.03 to 0.11) | 2,022,722 | 9 | 0.05 (0.02 to 0.09) | -0.01 (-0.06 to 0.04) |
| Prior SARS-CoV-2 infection | Yes | 10,969,377 | 15 | 0.03 (0.02 to 0.06) | 10,968,987 | 15 | 0.01 (0.01 to 0.02) | -0.02 (-0.04 to 0.00) |
|  | No | 606,015 | 0 | 0.00 (- to -) | 606,039 | 3 | 0.03 (0.01 to 0.11) | 0.03 (0.00 to 0.07) |
| JCVI clinical risk group | Not at-risk | 9,752,358 | 3 | 0.00 (0.00 to 0.01) | 9,751,968 | 9 | 0.01 (0.01 to 0.03) | 0.01 (0.00 to 0.02) |
|  | At-risk | 1,666,876 | 9 | 0.08 (0.04 to 0.15) | 1,666,776 | 9 | 0.06 (0.03 to 0.12) | -0.01 (-0.08 to 0.05) |
|  | Extremely vulnerable | 156,146 | 3 | 0.19 (0.06 to 0.59) | 156,360 | 3 | 0.19 (0.06 to 0.59) | 0.00 (-0.30 to 0.30) |

The 12-week risks (cumulative incidences) per 1,000 people of a positive SARS-CoV-2 test were 103.2 (95%CI 102.4 to 104.0) for those receiving BNT162b2 and 96.0 (95.2 to 96.8) for mRNA-1273 respectively, representing a risk difference of − 7.24 (95%CI − 8.37 to − 6.11) (Table 2). Corresponding 12-week risks of COVID-19 hospitalisation were 0.65 (0.56 to 0.75) and 0.44 (0.36 to 0.54) per 1,000 people (RD − 0.21 (− 0.34 to − 0.08)), and of COVID-19 death 0.03 (0.02 to 0.06) and 0.01 (0.01 to 0.02) per 1,000 people (RD − 0.02 (− 0.04 to 0.00)). Absolute risks differed between subgroups, and therefore so did the absolute risk differences comparing vaccines, although mRNA-1273 was consistently associated with a reduction in risk compared with BNT162b2 for positive SARS-CoV-2 tests and COVID-19 hospitalisation (Table 2).

The hazard ratios comparing mRNA-1273 with BNT162b2 were 0.92 (95%CI 0.91 to 0.92) for positive SARS-CoV-2 test, 0.67 (0.58 to 0.78) for COVID-19 hospitalisation, and 1.23 (0.59 to 2.56) for COVID-19 death (Figure 3). For positive SARS-CoV-2 tests the relative benefit of mRNA-1273 compared with BNT162b2 was somewhat greater in those who received BNT162b2 for their first two vaccine doses (HR 0.90 (0.89 to 0.91)) than in those who received ChAdOx1 (HR 0.93 (0.92 to 0.93), heterogeneity p=0.002). Similarly, the relative benefit of mRNA-1273 compared with BNT162b2 was greater in those with prior evidence of infection (HR 0.87 (0.83 to 0.91)) versus those without (HR 0.92 (0.91 to 0.93), heterogeneity p=0.010). For COVID-19 hospitalisation, hazard ratios were broadly similar across the different subgroups. For COVID-19 death, HRs were estimated imprecisely and so no strong conclusions about variation between subgroups could be drawn. Supplementary materials provide cumulative incidence plots by subgroup (Figures S3a-e), the corresponding risk differences and risk ratios over 12-weeks (Figures S4a-e), and subgroup-specific heterogeneity tests for risk differences and risk ratios (Table S1).

**Figure 3:**
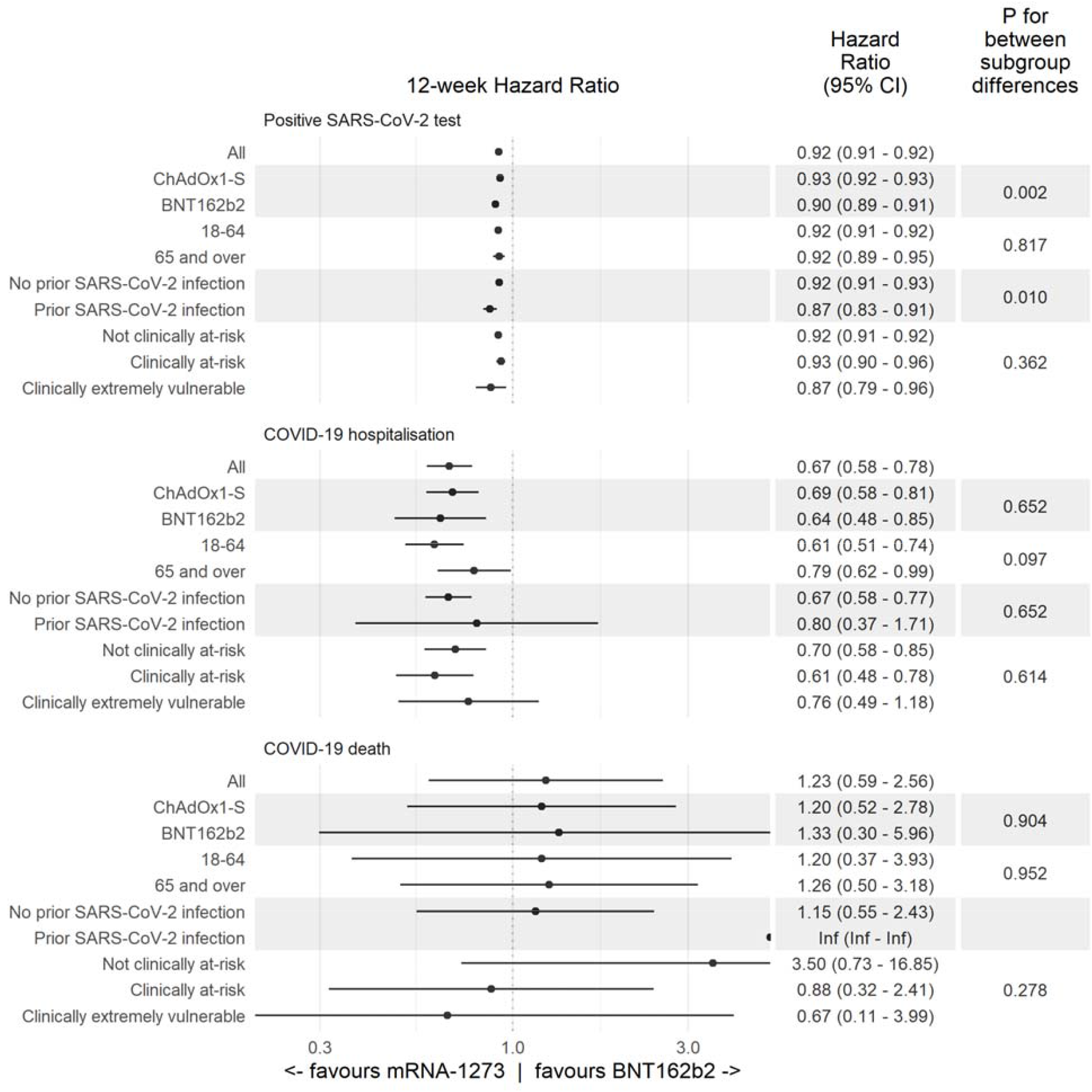
Estimated hazard ratios comparing the effectiveness of mRNA-1273 and BNT162b2, overall and within subgroups, together with p-values for between-subgroup heterogeneity

## Discussion

This observational study in over 3 million adults compared the effectiveness of BNT162b2 and mRNA-1273 booster vaccination and is, to our knowledge, the first to compare their effectiveness against severe COVID-19 outcomes. We estimated that mRNA-1273 provided better protection than BNT162b2 against a positive SARS-CoV-2 test (HR = 0.92 (95%CI 0.91 to 0.92)) and, particularly, COVID-19 hospitalisation (0.67 (95%CI 0.58 to 0.78)) in the first 12 weeks after boosting. There were only 30 COVID-19 deaths across both vaccine groups, so the hazard ratio estimated imprecisely.

Around 1 in 14 people had a positive SARS-CoV-2 test in the 12 weeks following booster vaccination reflecting both the high infection rates in the study period, particularly from December 2021 onward as the Omicron variant become dominant, and the imperfect protection against infection offered by booster vaccination. The relative advantage of mRNA-1273 over BNT162b2 in preventing positive SARS-CoV-2 tests was greater in people whose primary vaccination course was BNT162b2 compared with ChAdOx1, suggesting a possible benefit of heterologous boosting in preventing infection.

Severe COVID-19 disease was rare up to 12-weeks after booster vaccination: fewer than 1 in 3,800 booster recipients were hospitalised with COVID-19, and fewer than 1 in 100,000 died with COVID-19 as an underlying or contributory cause. The relative effectiveness of mRNA-1273 versus BNT162b2 against COVID-19 hospitalisation appeared similar regardless of age, clinical vulnerability, and whether there was evidence of prior infection.

### Strengths and limitations

The OpenSAFELY-TPP database covers around 40% of the English population and contains rich clinical information which enabled us to closely match BNT162b2 and mRNA-1273 recipients to control for potential confounding, to study a range of clinical outcomes, including severe COVID-19, and compare comparative effectiveness between important clinical subgroups.

To make fair comparisons between the two vaccine types, we exploited the concurrent roll-out of both vaccines across the same eligible population in the same time period. The type of vaccine administered was based largely on local supply and availability, and clinical characteristics did not inform the type of vaccine offered on-site. It remained important to control for any potential differences in the distribution of prognostic factors between the two vaccine groups, and these were well-balanced after matching. We cannot rule out residual confounding, but unmeasured confounding, particularly relating to unmeasured health-seeking behaviours, are less problematic in comparative effectiveness studies than in studies comparing vaccinated with unvaccinated people, as all people entering the study have sought and received a booster dose.

We excluded groups who were unlikely to undergo boosting such as those with a recent positive test and in palliative care. We also excluded groups where we could not reliably capture heterogeneity in outcome risk such as those living in care homes and health and social care workers. The later introduction of mRNA-1273 booster vaccination meant that the matched cohort contained younger and healthier people than the whole boosted population. These restrictions may limit the generalisability of our findings. However, numbers remained sufficient to compare subgroups defined by age, clinical vulnerability, and prior infection status, including for COVID-19 hospitalisation.

To control for spatio-temporal heterogeneity in infection risk during the study period we matched on the date of booster vaccine receipt and local health administration (STP). Some residual heterogeneity within STP regions is possible, but more precise geographical matching was not feasible due to poor matching success at this level, partly due to time periods during which only one vaccine type was available within a small geographical area.

Some outcomes may be under-ascertained. Positive SARS-CoV-2 tests only include those reported via the national COVID-19 surveillance system (SGSS), and so many asymptomatic and some symptomatic infections will have been missed. Some COVID-19 hospitalisations resulting in longer hospital stays may have been omitted from analyses as these events are only recorded in the data available within OpenSAFELY once the patient has been discharged.

This study does not include anyone who received mRNA-1273 as a primary course where, due to potential benefits of heterologous vaccination, boosting with BNT162b2 may have outperformed mRNA-1273. However, subgroup analyses in those who received ChAdOx1 as a primary course, therefore where all subsequent boosting was heterologous, demonstrated that the estimated superiority of mRNA-1273 remained.

### Findings in context

The COV-BOOST phase II randomised trial assessed the safety and immunogenicity of seven COVID-19 vaccines for boosting, including BNT162b2 and mRNA-1273 against a menACWY control in people with no prior history of infection (7). It found higher immunogenicity of mRNA-1273 compared with BNT162b2 in subgroups with a two-dose BNT162b2 primary course (geometric mean ratio (GMR) of SARS-CoV-2 anti-spike IgG versus control = 11.49 (95%CI 9.36 to 14.12) versus (6.78 (5.51 to 8.35)) and a two-dose ChAdOx1 primary course (GMR = 32.30 (24.84 to 42.01) versus 16.80 (12.97 to 21.76)). Likewise, the MixNMatch phase I-II non-randomised trial documented higher antibody levels after mRNA-1273 boost than BNT162b2 boost among individuals who received a primary course of BNT162b2 (8). These trials did not evaluate efficacy endpoints, and we are aware of no other planned or published randomised trials making direct comparisons of boosting with BNT162b2 versus mRNA-1273.

A study in Spanish registry data of people aged 40 years or over with no previous positive test for SARS-CoV-2 examined effectiveness of mRNA vaccine boosting, including a comparison of BNT162b2 with mRNA-1273 (9). For positive SARS-CoV-2 tests, it estimated a 34-day risk difference comparing mRNA-1273 with BNT162b2 of − 2.2 (− 2.7 to − 1.6) per 1,000 people (risk ratio 0.87 (0.84, 0.90)). There was no assessment of effectiveness against hospitalisation or other severe outcomes.

We are aware of no other observational studies comparing effectiveness for booster vaccination of BNT162b2 and mRNA-1273 in the context of booster vaccination, though comparisons have been made for primary vaccination. A study using data from the US Veteran Affairs health care system during the Alpha period using a similar matching approach to the present study (10), identified an additional 1.23 (95% CI 0.72 to 1.81) documented infections per 1,000 people in those with a BNT162b2 first dose compared with a mRNA-1273 first dose at 24 weeks, 0.55 (95%CI 0.36 to 0.83) COVID-19 hospitalisations, 0.10 (95%CI 0.00 to 0.26) COVID-19 ICU admissions, and 0.02 (95%CI − 0.06 to 0.12) COVID-19 deaths. Another study using Veterans Affairs data with exact- and propensity-matching in a similar time period (11), estimated similar differences at 24 weeks for documented infection (1.73, 95%CI 1.50 to 1.96 additional events per 1,000 people in those with a BNT162b2 first dose compared with a mRNA-1273), COVID-19 hospitalisation (0.56, 95%CI 0.45 to 0.67) and COVID-19 death (0.03, 95%CI -0.00 to 0.07). Accounting for the longer follow-up period and statistical uncertainty, these estimates are consistent with our findings for the comparative effectiveness of mRNA-1273 versus BNT162b2 boosting.

A study using English data to estimate the effectiveness of BNT162b2 boosting versus no boosting and mRNA-1273 boosting versus no boosting separately using a test-negative-control design (12), suggests marginally higher effectiveness against symptomatic disease in mRNA-1273 than in BNT162b2, but this assumes that the respective control populations in each analysis were similar.

The evidence therefore strongly points to a benefit of mRNA-1273 over BNT162b2 for primary vaccination and subsequent booster doses. This is relevant to vaccine procurement decisions for future booster programmes. However, both vaccines are safe (13–16) and strongly effective (9,12,17,18) against infection and COVID-19 disease, compared to no boosting. Findings from this study should not discourage individuals from receiving BNT162b2 booster vaccination if offered.

## Data Availability

Detailed pseudonymised patient data is potentially re-identifiable and therefore not shared. All study code is available for inspection and re-use at https://github.com/opensafely/comparative-booster

## Contributions

Conceptualization: WJH, EMFH, EPKP, RHK, EJW, VW, TP, BG, MAH, and JACS. Data curation: WJH, HJC, AW, AM, BM, and SCJB. Formal analysis: WJH, EMFH, EPKP, and VW. Funding acquisition: JM, BG, and JACS. Methodology: WJH, EMFH, EPKP, RHK, EJW, VW, TP, MAH, and JACS. Project administration: WJH, AM, JM, BG, and JACS. Resources: WJH, HJC, AW, AM, JM, BM, SCJB, and BG. Software: WJH, EMFH, EPKP, and SCJB. Supervision: MAH and JACS. Validation: WJH, EMFH, EPKP, and VW. Visualization: WJH, EMFH, and EPKP. Writing - original draft: WJH. Writing - review & editing: WJH, EMFH, EPKP, RHK, EJW, VW, TP, HJC, AW, AM, JM, BM, SCJB, BG, MAH, and JACS.

### Information governance and ethical approval

NHS England is the data controller for OpenSAFELY-TPP; TPP is the data processor; and study authors using OpenSAFELY have the approval of NHS England. This implementation of OpenSAFELY is hosted within the TPP environment which is accredited to the ISO 27001 information security standard and is NHS IG Toolkit compliant (19); Patient data has been pseudonymised for analysis and linkage using industry standard cryptographic hashing techniques; all pseudonymised datasets transmitted for linkage onto OpenSAFELY are encrypted; access to the platform is via a virtual private network (VPN) connection, restricted to a small group of researchers; the researchers hold contracts with NHS England and only access the platform to initiate database queries and statistical models; all database activity is logged; only aggregate statistical outputs leave the platform environment following best practice for anonymisation of results such as statistical disclosure control for low cell counts (20). The OpenSAFELY research platform adheres to the obligations of the UK General Data Protection Regulation (GDPR) and the Data Protection Act 2018. In March 2020, the Secretary of State for Health and Social Care used powers under the UK Health Service (Control of Patient Information) Regulations 2002 (COPI) to require organisations to process confidential patient information for the purposes of protecting public health, providing healthcare services to the public and monitoring and managing the COVID-19 outbreak and incidents of exposure; this sets aside the requirement for patient consent (21). Taken together, these provide the legal bases to link patient datasets on the OpenSAFELY platform. General practices, from which the primary care data are obtained, are required to share relevant health information to support the public health response to the pandemic, and have been informed of the OpenSAFELY analytics platform.

This study was approved by the Health Research Authority (REC reference 20/LO/0651) and by the LSHTM Ethics Board (reference 21863).

JACS is the guarantor.

## Data access and verification

Access to the underlying identifiable and potentially re-identifiable pseudonymised electronic health record data is tightly governed by various legislative and regulatory frameworks, and restricted by best practice. The data in OpenSAFELY is drawn from General Practice data across England where TPP is the data processor.

TPP developers initiate an automated process to create pseudonymised records in the core OpenSAFELY database, which are copies of key structured data tables in the identifiable records. These pseudonymised records are linked onto key external data resources that have also been pseudonymised via SHA-512 one-way hashing of NHS numbers using a shared salt. Bennett Institute for Applied Data Science developers and PIs holding contracts with NHS England have access to the OpenSAFELY pseudonymised data tables as needed to develop the OpenSAFELY tools.

These tools in turn enable researchers with OpenSAFELY data access agreements to write and execute code for data management and data analysis without direct access to the underlying raw pseudonymised patient data, and to review the outputs of this code. All code for the full data management pipeline—from raw data to completed results for this analysis—and for the OpenSAFELY platform as a whole is available for review at github.com/OpenSAFELY.

