## Supplementary material for "Comparative effectiveness of BNT162b2 versus mRNA-1273 boosting in England: a cohort study in OpenSAFELY-TPP": supplement.html

Supplementary materials for Comparative effectiveness of BNT162b2 versus mRNA-1273 booster doses in England


### Supplementary materials for Comparative effectiveness of BNT162b2 versus mRNA-1273 booster doses in England

###### Figure S1: Cumulative numbers included in the matched sample

###### Figure S2: Standardised Mean Differences between BNT12b2 and mRNA-1273 groups, before and after matching

###### Table S1: 12-week comparative effectiveness, with p-values for subgroup heterogeneity

| Sub-group |  | Risk difference per 1,000 people (95% CI) | P-value | Risk ratio (95% CI) | P-value | Hazard ratio (95% CI) | P-value |
| --- | --- | --- | --- | --- | --- | --- | --- |
| Positive SARS-CoV-2 test | | | | | | | |
| Main |  | −7.24 (−8.37 to −6.11) | – | 0.93 (0.92-0.94) | – | 0.92 (0.91 to 0.92) | – |
| Primary vaccine course | ChAdOx1-S | −5.39 (−6.68 to −4.10) | <0.001 | 0.94 (0.93-0.96) | 0.002 | 0.93 (0.92 to 0.93) | 0.002 |
|  | BNT162b2 | −11.66 (−14.09 to −9.24) | – | 0.91 (0.89-0.93) | – | 0.90 (0.89 to 0.91) | – |
| Age | 18-64 | −6.53 (−9.04 to −4.02) | 0.010 | 0.95 (0.93-0.97) | 0.286 | 0.92 (0.91 to 0.92) | 0.817 |
|  | 65 and over | −2.75 (−4.11 to −1.39) | – | 0.92 (0.89-0.96) | – | 0.92 (0.89 to 0.95) | – |
| Prior SARS-CoV-2 infection status | No prior SARS-CoV-2 infection | −7.21 (−8.36 to −6.06) | 0.746 | 0.93 (0.92-0.94) | 0.241 | 0.92 (0.91 to 0.93) | 0.010 |
|  | Prior SARS-CoV-2 infection | −8.09 (−13.26 to −2.91) | – | 0.89 (0.83-0.96) | – | 0.87 (0.83 to 0.91) | – |
| Clinical vulnerability | Not clinically at-risk | −7.44 (−8.86 to −6.02) | 0.045 | 0.93 (0.92-0.94) | 0.382 | 0.92 (0.91 to 0.92) | 0.362 |
|  | Clinically at-risk | −4.36 (−6.48 to −2.25) | – | 0.94 (0.91-0.97) | – | 0.93 (0.90 to 0.96) | – |
|  | Clinically extremely vulnerable | −9.10 (−15.55 to −2.66) | – | 0.87 (0.79-0.96) | – | 0.87 (0.79 to 0.96) | – |
| COVID-19 hospitalisation | | | | | | | |
| Main |  | −0.21 (−0.34 to −0.08) | – | 0.68 (0.53-0.87) | – | 0.67 (0.58 to 0.78) | – |
| Primary vaccine course | ChAdOx1-S | −0.05 (−0.25 to 0.15) | 0.003 | 0.92 (0.66-1.29) | 0.001 | 0.69 (0.58 to 0.81) | 0.652 |
|  | BNT162b2 | −0.64 (−0.98 to −0.30) | – | 0.37 (0.24-0.58) | – | 0.64 (0.48 to 0.85) | – |
| Age | 18-64 | −0.14 (−0.26 to −0.01) | 0.665 | 0.70 (0.49-0.99) | 0.500 | 0.61 (0.51 to 0.74) | 0.097 |
|  | 65 and over | −0.19 (−0.42 to 0.03) | – | 0.81 (0.63-1.04) | – | 0.79 (0.62 to 0.99) | – |
| Prior SARS-CoV-2 infection status | No prior SARS-CoV-2 infection | −0.30 (−0.47 to −0.14) | 0.343 | 0.60 (0.45-0.78) | 0.271 | 0.67 (0.58 to 0.77) | 0.652 |
|  | Prior SARS-CoV-2 infection | −0.19 (−0.36 to −0.01) | – | 0.36 (0.15-0.85) | – | 0.80 (0.37 to 1.71) | – |
| Clinical vulnerability | Not clinically at-risk | 0.07 (−0.11 to 0.24) | <0.001 | 1.19 (0.77-1.85) | 0.017 | 0.70 (0.58 to 0.85) | 0.614 |
|  | Clinically at-risk | −0.52 (−0.77 to −0.28) | – | 0.57 (0.44-0.74) | – | 0.61 (0.48 to 0.78) | – |
|  | Clinically extremely vulnerable | −0.93 (−2.30 to 0.44) | – | 0.73 (0.46-1.16) | – | 0.76 (0.49 to 1.18) | – |
| COVID-19 death | | | | | | | |
| Main |  | −0.02 (−0.04 to 0.00) | – | 0.35 (0.16-0.77) | – | 1.23 (0.59 to 2.56) | – |
| Primary vaccine course | ChAdOx1-S | 0.00 (−0.01 to 0.01) | 0.202 | 0.79 (0.31-2.00) | 0.110 | 1.20 (0.52 to 2.78) | 0.904 |
|  | BNT162b2 | −0.03 (−0.08 to 0.01) | – | 0.17 (0.04-0.86) | – | 1.33 (0.30 to 5.96) | – |
| Age | 18-64 | 0.00 (0.00 to 0.00) | 0.709 | 0.99 (0.20-4.90) | 0.858 | 1.20 (0.37 to 3.93) | 0.952 |
|  | 65 and over | −0.01 (−0.06 to 0.04) | – | 0.84 (0.33-2.11) | – | 1.26 (0.50 to 3.18) | – |
| Prior SARS-CoV-2 infection status | No prior SARS-CoV-2 infection | −0.02 (−0.04 to 0.00) | 0.015 | 0.39 (0.17-0.85) | – | 1.15 (0.55 to 2.43) | <0.001 |
|  | Prior SARS-CoV-2 infection | 0.03 (0.00 to 0.07) | – | Inf (NA-NA) | – | Inf (Inf to Inf) | – |
| Clinical vulnerability | Not clinically at-risk | 0.01 (0.00 to 0.02) | 0.746 | 6.48 (1.71-24.50) | 0.039 | 3.50 (0.73 to 16.85) | 0.278 |
|  | Clinically at-risk | −0.01 (−0.08 to 0.05) | – | 0.82 (0.32-2.11) | – | 0.88 (0.32 to 2.41) | – |
|  | Clinically extremely vulnerable | 0.00 (−0.30 to 0.30) | – | 1.00 (0.20-4.94) | – | 0.67 (0.11 to 3.99) | – |

##### Cumulative incidence


###### Figure S3a: Cumulative incidence estimates per 1,000 people

###### Figure S3b: Cumulative incidence estimates per 1,000 people by primary course type

###### Figure S3c: Cumulative incidence estimates per 1,000 people for by age group

###### Figure S3d: Cumulative incidence estimates per 1,000 people by prior infection status

###### Figure S3e: Cumulative incidence estimates per 1,000 people by clinical vulnerability

##### Cumulative contrasts

###### Figure S4a: Cumulative risk difference and risk ratio

###### Figure S4b: Cumulative risk difference and risk ratio by primary course type

###### Figure S4c: Cumulative risk difference and risk ratio by age group

###### Figure S4d: Cumulative risk difference and risk ratio by prior infection status

###### Figure S4e: Cumulative risk difference and risk ratio by clinical vulnerability
